# Vaccination strategies when vaccines are scarce: On conflicts between reducing the burden and avoiding the evolution of escape mutants

**DOI:** 10.1101/2021.05.04.21256623

**Authors:** Félix Geoffroy, Arne Traulsen, Hildegard Uecker

## Abstract

When vaccine supply is limited but population immunisation urgent, the allocation of the available doses needs to be carefully considered. One aspect of dose allocation is the time interval between the primer and the booster injections in two-dose vaccines. By stretching this interval, more individuals can be vaccinated with the first dose more quickly. Even if the level of immunity of these ‘half-vaccinated’ individuals is lower than that of those who have received both shots, delaying the second injection can be beneficial in reducing case numbers, provided a single dose is sufficiently effective. On the other hand, there has been concern that intermediate levels of immunity in partially vaccinated individuals may favour the evolution of vaccine escape mutants. In that case, a large fraction of half-vaccinated individuals would pose a risk – but only if they encounter the virus. This raises the question whether there is a conflict between reducing the burden and the risk of vaccine escape evolution or not. We develop a minimal model to assess the population-level effects of the timing of the booster dose. We set up an SIR-type model, in which more and more individuals become vaccinated with a two-dose vaccine over the course of a pandemic. As expected, there is no trade-off when vaccine escape evolves at equal probabilities in unvaccinated and half-vaccinated patients. If vaccine escape evolves more easily in half-vaccinated patients, the presence or absence of a trade-off depends on the reductions in susceptibility and transmissibility elicited by the primer dose.

## Introduction

Many vaccines are administered in two doses, a primer shot and – after a certain time interval – a booster shot. The booster shot increases the strength and duration of protection. However, the primer shot on its own already establishes some immunity. In a pandemic – such as in the current Covid pandemic – when population immunisation is urgent but vaccine doses are scarce, the question arises whether the second shot should be delayed at the benefit of administering the first vaccine dose to more people more quickly. Even if half-vaccinated individuals are only partially immune, the overall reduction in infections may be greater in a population in which many people have some immunity than in a population in which fewer individuals have stronger immunity. In the current Covid pandemic, such a delay strategy has been adopted by the UK (Campbell, 2020 Dec 30), while several other countries such as the US stick to the interval between injections that has been applied in the original clinical trials and is therefore recommended by the manufacturer (U.S. Food and Drug Administration). In early January 2021, the WHO recommended to stretch the dosing interval of the first approved vaccine (Pfizer–BioNTech COVID-19 vaccine) from 21-28 days to 42 days in countries ‘experiencing exceptional epidemiological circumstances’ (World Health Organization,2021), which has for example been adopted by Germany, especially from April on (Vygen-Bonnet et al.,2021). Already preceding the current pandemic, mathematical models have compared the effects of a delay strategy in its extreme form – a one-dose strategy – and a two-dose strategy for cholera and influenza epidemics/pandemics (Azman et al.,2015; Matrajt et al., 2015). Sparked by the Covid crisis, a series of models have been set up to assess when stretching the period between the primer and booster shots reduces the total number of SARS-CoV-2 infections (Böttcher and Nagler, 2021;Kraay et al., 2021; Matrajt et al., 2021; Paltiel et al.,2021; Saad-Roy et al.,2021).

However, there is another dimension to the problem, since the vaccination strategy does not only affect the dynamics of the pandemic, but may also influence the evolutionary dynamics of the virus (or another pathogen in other circumstances). This especially concerns the evolution of vaccine escape mutants against which the vaccine has no or reduced efficiency (Saad-Roy et al., 2021). Vaccine resistance is generally rare (Kennedy and Read,2017). Yet, the large case numbers in the current pandemic give the virus a lot of opportunity to replicate, mutate, and adapt. It has been hypothesized that vaccine escape mutants evolve most easily in people who have only received one vaccine shot (Branswell,2021 Jan. 4). The reasoning behind this hypothesis is the following (similar to Grenfell et al.,2004): After the primer dose, vaccinees have intermediate levels of antibodies (Krammer et al., 2021;Saad-Roy et al.,2021). This level is not enough to keep the viral load following exposure to the virus sufficiently low to avoid the occurrence of a large number of mutations, including vaccine escape mutations. At the same time, it gives a large advantage to mutant virus particles to which the antibodies bind only weakly. In contrast, in fully vaccinated individuals, the viral load is kept low such that mutations are unlikely to occur. In unvaccinated patients, the virus can initially replicate well and attain high numbers. However, the immune response is broader than the one elicited by the vaccine such that vaccine escape mutants do not have a great advantage over other viral genotypes. Hence, according to this reasoning, the evolution of vaccine escape is most likely at intermediate levels of antibodies, which are typical for half-vaccinated individuals (but see the counter-arguments made by Cobey et al.,2021). This raises the concern that the large number of half-vaccinated individuals in the delay strategy may drive the evolution of vaccine escape.

Viral evolution can only occur in infected individuals. Thus, even if the above reasoning holds, whether vaccine escape evolves in a host population does not only depend on the number of partially immune individuals, but also on the number of infected individuals. If the delay strategy reduces the disease prevalence in the population, this may offset the increased probability of vaccine escape evolution within any one half-vaccinated patient and may actually decrease rather than increase the risk of vaccine escape (see the discussion in Cobey et al.,2021). Hence, in the interplay of all effects, is there a trade-off between reducing the cumulative number of infections in the pandemic and minimising the risk of escape mutants or not?

What we were missing in the current public debate is a quantitative epidemic model that includes the emergence of escape mutations and quantifies how the strengths of the various population-level effects compare to each other. We therefore set up an SIR-type model to dissect and quantify the considerations and verbal arguments outlined above. We chose a minimal model that is stripped down to the most essential components needed to study both aspects of the problem of dose allocation. This, of course, ignores much of the biological complexity and does not allow to make immediate recommendations for vaccine strategies in the current pandemic. However, the transparency of the model makes it possible to develop a better intuition for the conditions under which there is a trade-off and those under which the same allocation strategy is optimal in both respects. We therefore hope that it can contribute to a better-informed discussion.

## The Model

We consider an SIR-type model, where individuals are either unvaccinated, vaccinated with the primer dose only, or fully vaccinated with both doses. Since we are interested in the rate of *de novo* emergence of vaccine escape mutants and not in their subsequent spread, we model the disease dynamics in the absence of the vaccine escape variant. From the number of wild-type infections, we can estimate the risk of vaccine escape evolution. The flow diagram of the model is shown in Fig. 1a.

**Figure 1:**
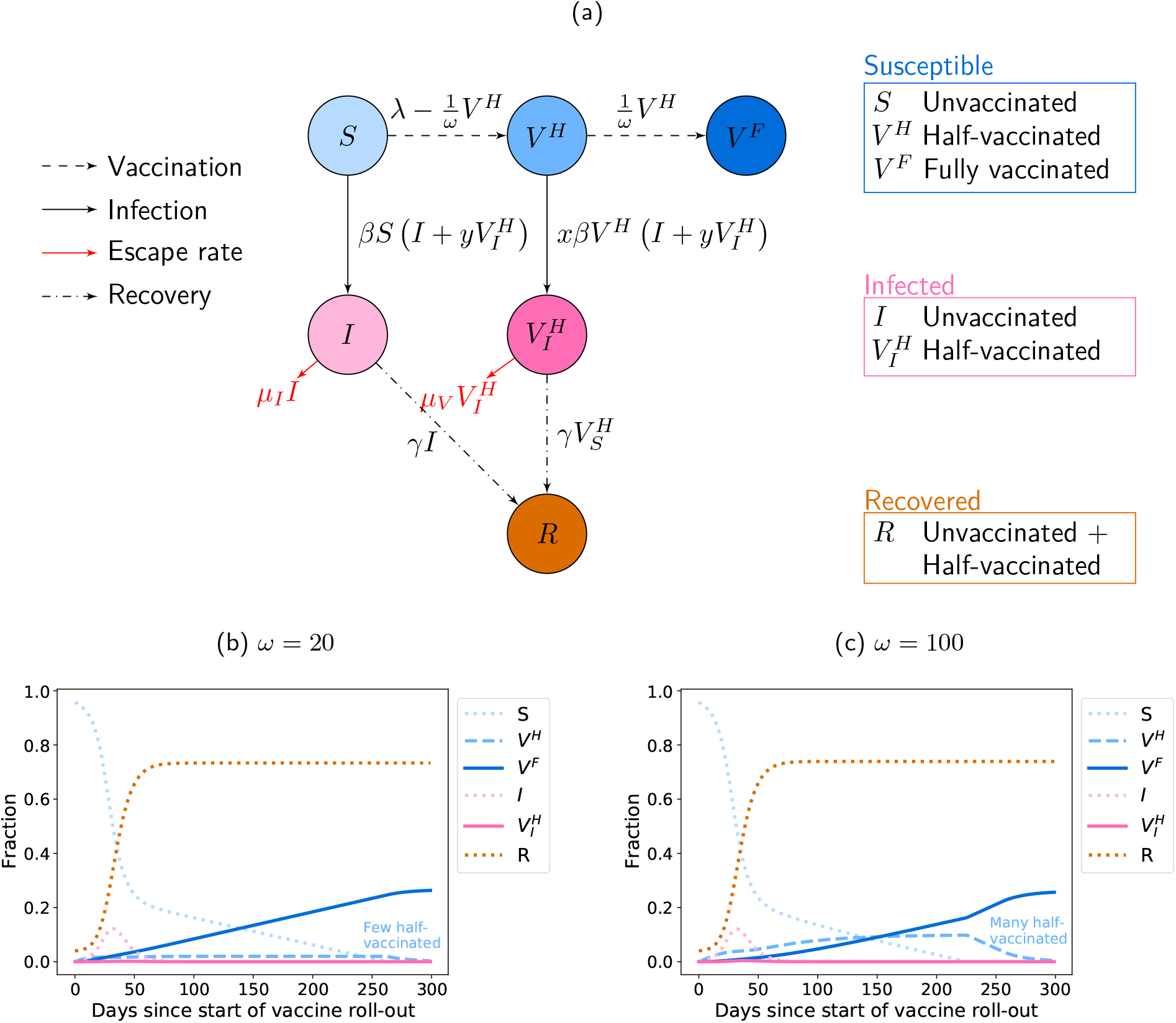
Model for the disease dynamics with a two-dose vaccine. (a) Flow diagram of the model defined by Eq. (1). The model corresponds to an extended SIR model, in which individuals can either be unvaccinated, vaccinated with the primer dose only, or vaccinated with the primer and booster doses. The primer dose reduces susceptibility by a factor *x* and transmissibility by a factor *y*. Vaccination with both doses provides perfect protection from the virus. Since vaccine supply is limited, there is a trade-off: either administering the booster shots after the minimally required prime-boost interval *ω*_min_ or delaying the booster injection and giving the primer dose to more people more quickly (*ω > ω*_min_). The model describes the dynamics in the absence of vaccine escape mutants. The rate at which such mutants emerge can be obtained from the number of infected individuals, where vaccine escape may possibly evolve at different per-capita rates in unvaccinated and half-vaccinated patients. The allocation strategy affects the total number of individuals that become infected throughout the pandemic, but also the risk of vaccine escape evolution across the population. It is apriori not clear whether the same strategy minimises both quantitites. (b-c) Exemplary dynamics of infection and vaccination with and without a delay in the booster dose. In Panel (b), the booster dose is administered as soon as possible (*ω* = *ω*_min_). In this case, the fraction of half-vaccinated individuals is always low. In Panel (c), the booster dose is delayed (*ω* = *ω*_max_), leading to a much higher fraction of half-vaccinated individuals over time. The vaccination campaign starts during the pandemic, when a considerable fraction of the population has already been affected by the virus (*I*(0) = 2 *×* 10^*-*3^ and *R*(0) = 4 *×* 10^*-*2^). For illustrative purposes, we use a large reproductive number *R*_*C*_ = 2 here.

Once vaccines become available, there is a limited but constant supply of vaccine doses that allows to administer injections at total rate *λ*. We assume that vaccine doses are only given to individuals who have never been infected by the virus (i.e. infected and recovered individuals do not receive any (further) vaccination). Any vaccine dose can either be used as a primer or as a booster shot. If the time interval between the two injections is chosen as *ω* and the booster shots are administered at rate 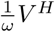, the rate of primer dose injections is limited to 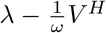. The time interval *ω* determines the vaccination strategy: it can either be set to the interval *ω*_min_ recommended by the manufacturer or be stretched to increase the rate at which unvaccinated individuals receive the first dose. We set the maximal interval as *ω*_max_ = 5 *ω*_min_. For simplicity, we assume that the efficiency of the primer dose remains constant over time and that the dosing interval *ω* has no effect on the efficiency of the booster dose.

Unvaccinated and partially vaccinated individuals can become infected with the virus. Vaccination with both doses, however, entirely blocks infection with the wild-type strain. The transmission coefficient between unvaccinated infected individuals and unvaccinated susceptible individuals is given by *β*. Vaccination with the primer dose reduces susceptibility by a factor *x* and transmissibility by a factor *y* (Gandon and Day,2007;Matrajt et al., 2015). We do not explicitly model any vaccine effect on pathogenicity (unlike Gandon and Day (2007);Matrajt et al.(2015)), and we assume that infected individuals recover at rate *γ*, irrespective of their vaccination status. Upon recovery, all infected individuals gain full immunity over the relevant time scales of the pandemic. We assume that vaccine escape variants against which the vaccine has reduced efficiency evolve and become dominant within unvaccinated individuals at per-capita rate *µ*_*I*_ and within half-vaccinated individuals at per-capita rate *µ*_*V*_ *≥ µ*_*I*_. Once the vaccine escape variant is dominant in one individual, it can spread across the entire population. However, here we focus on the emergence of these strains and not on their future dynamics.

Before the escape variant arises, the dynamics of the epidemic is described by the differential equations:

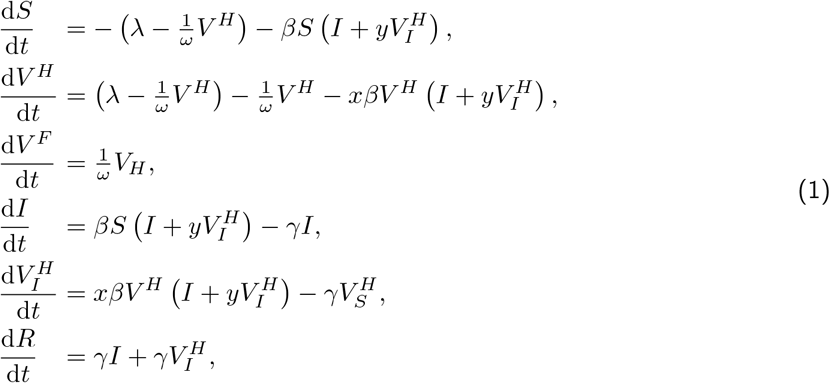

where the first three equations describe individuals that are uninfected and either unvaccinated (*S*), half-vaccinated (*V* ^*H*^), or fully vaccinated (*V* ^*F*^). The next two equations describe individuals that are infected and either unvaccinated (*I*) or half-vaccinated 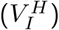. The last equation describes all individuals that are immune due to naturally acquired immunity following an infection (*R*).

Once every individual has received the primer dose (or has been infected by the virus), i.e. *S* = 0, there is no reason anymore to delay the booster dose. However, it is possible that in a delay strategy, many half-vaccinated individuals are waiting for the second shot and that the available doses are still not sufficient to vaccinate them at rate 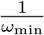. Therefore, once *S* = 0, the second shot is administered at a rate given by the minimum of 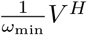 and *λ*. (As long as *S >* 0, there are always enough doses available to vaccinate at a steady per-capita rate 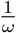, see Appendix A.) Ultimately, every individual in the population has either been vaccinated or has acquired immunity through infection.

We choose the model parameters in accordance with values for the Covid pandemic. We set the infectious period to 5 days, i.e.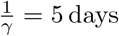 days (cf. Saad-Roy et al.,2021) and the minimal interval between the vaccine doses to *ω*_min_ = 20 days, which is the recommended interval for the Biontech/Pfizer vaccine. For the rate of vaccine roll-out, if not stated otherwise, we choose *λ* = 0.2%, which roughly corresponds to the rate in Germany during March 2021 (Ritchie et al.).

As a default, we assume the reproductive number in the absence of any immune individuals to be 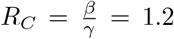, which is much smaller than the basic reproductive number of SARS-CoV-2 in the absence of control measures (Bar-On et al.,2020). Our choice means that there are control measures in place, but they are insufficient to control the spread of the disease. There are no estimates for the per-patient mutation rates *µ*_*I*_ and *µ*_*V*_. The absolute values only change the number of mutant infections, while the qualitative results in our model depend on the ratio *µ*_*V*_ */µ*_*I*_ (see below for details). To account for the uncertainty in this ratio, we consider *µ*_*V*_ */µ*_*I*_ = 1, 10, 100. We set *µ*_*I*_ = 10^*-*6^. Our primary focus is a scenario in which the vaccination campaign starts several months into the pandemic when a noticeable fraction of the population has already been affected by the virus. We define *t* = 0 as the start of the vaccination campaign. As initial conditions, we then set *I*(0) = 2 *×* 10^*-*3^ and *R*(0) = 4 *×* 10^*-*2^ (Roser et al., 2020), which corresponds approximately to twice the number of confirmed cases in Germany in January with the assumption that only 50% of the cases are detected (Backhaus et al.,2021).

We numerically integrate the differential equations using python, see our Jupyter notebook that is available with this manuscript. Exemplary dynamics are shown in Fig. 1b and c. In Panel (b), the booster dose it given as soon as possible (*ω* = *ω*_min_), while Panel (c) shows the dynamics under the maximal delay strategy (*ω* = *ω*_max_).

We aim to determine how the strategy affects the number of cases and the risk of vaccine escape. The first quantity that we consider is therefore the cumulative fraction of infected individuals from the start of the vaccination campaign until the end of the pandemic, provided no vaccine escape mutants evolve:

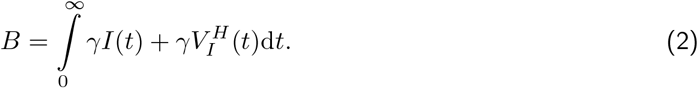

We refer to this as the burden. The total burden can be decomposed into the burden 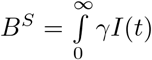 arising from unvaccinated individuals and the burden 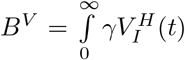 arising from half-vaccinated individuals. If the primer dose reduces the severity of disease, it is especially important to reduce *B*^*S*^, since this would reduce the number of severe cases.

The second quantity of interest is the total fraction of patients in whom vaccine escape mutants evolve after vaccine roll-out has started in the population (assuming that none are spreading yet at that time):

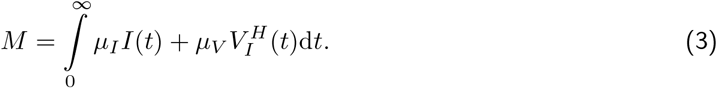

As for the burden, it can be insightful to decompose *M* according to the vaccination status of the patients into 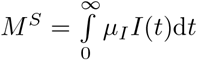 and 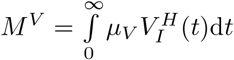.

In a non-deterministic world, these mutants may or may not evolve. The sum 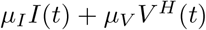 can also be interpreted as a stochastic rate, and the integral quantifies the total risk over the course of the pandemic (not taking into account the risk prior to the vaccine roll-out, which is independent of the chosen vaccination strategy). More precisely, the probability that vaccine escape mutants evolve is given by

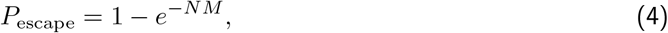

where *N* is the total population size. It should be noted that this is the probability of their mere appearance. That does not mean that they will spread. E.g., some of those patients in whom vaccine escape mutants evolve may not infect anyone, in which case the mutation is lost again from the population. For shortness, we often refer to *M* as the risk of vaccine escape in the following, but it should be kept in mind that the probability of vaccine escape is not directly given by *M* but by Eq. (4) and that this does not involve any probability of establishment of the vaccine escape mutant in the population.

## Results

### When can conflicts between reducing the burden and the escape risk appear?

What is the relationship between the burden *B* and the fraction of new mutant infections *M* ? The fraction of mutants can be rewritten as 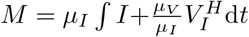. Thus, the qualitative effect of increasing or decreasing *ω* only depends on the ratio *µ*_*V*_ */µ*_*I*_, but not on their individual values. Comparing to the total burden 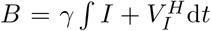, we see that there is no conflict between minimising both quantities if *µ*_*V*_ = *µ*_*I*_, since in that case both quantities are proportional to each other, *M ∝ B*. However, if *µ*_*V*_ *> µ*_*I*_, the two quantities are no longer proportional to each other, and it is conceivable that *M* increases with a change in *ω*, while *B* decreases. In contrast, we always have *M*^*S*^ *∝ B*^*S*^ and *M*^*V*^ *∝ B*^*V*^, i.e. within the two sub-populations of susceptibles and half-vaccinated patients there is no conflict and a reduction in burden always implies a reduction in escape risk.

### How do reductions in either susceptibility or transmissibility affect the burden and the risk of vaccine escape?

Before exploring the entire range of possible effect sizes *x* and *y* of the first vaccine dose, we consider the two limiting cases, in which the first vaccine dose has either an effect on susceptibility only (Fig. 2a-f) or on transmissibility only (Fig. 2g-l), but not on both simultaneously. For our default parameter set, reductions in susceptibility and transmissibility reduce both the burden and the escape risk, and they can both change the effect of the delay between the injections. With respect to the total burden, reductions in susceptibilty and transmissibility have overall very similar effects, both quantitatively and with respect to their influence on the optimal strategy (compare Panels a and g, but see results for higher *R*_*C*_ in Fig. 3, where this does not hold). In contrast, for the appearance of vaccine escape variants, it makes a difference whether the primer dose reduces susceptibility or transmissibilty (compare Panels d and j). The discrepancy comes from slight differences in the fraction of half-vaccinated patients that do not affect the total burden much but amplify in *M*^*V*^ (see especially the solid lines in Panels f and l).

**Figure 2:**
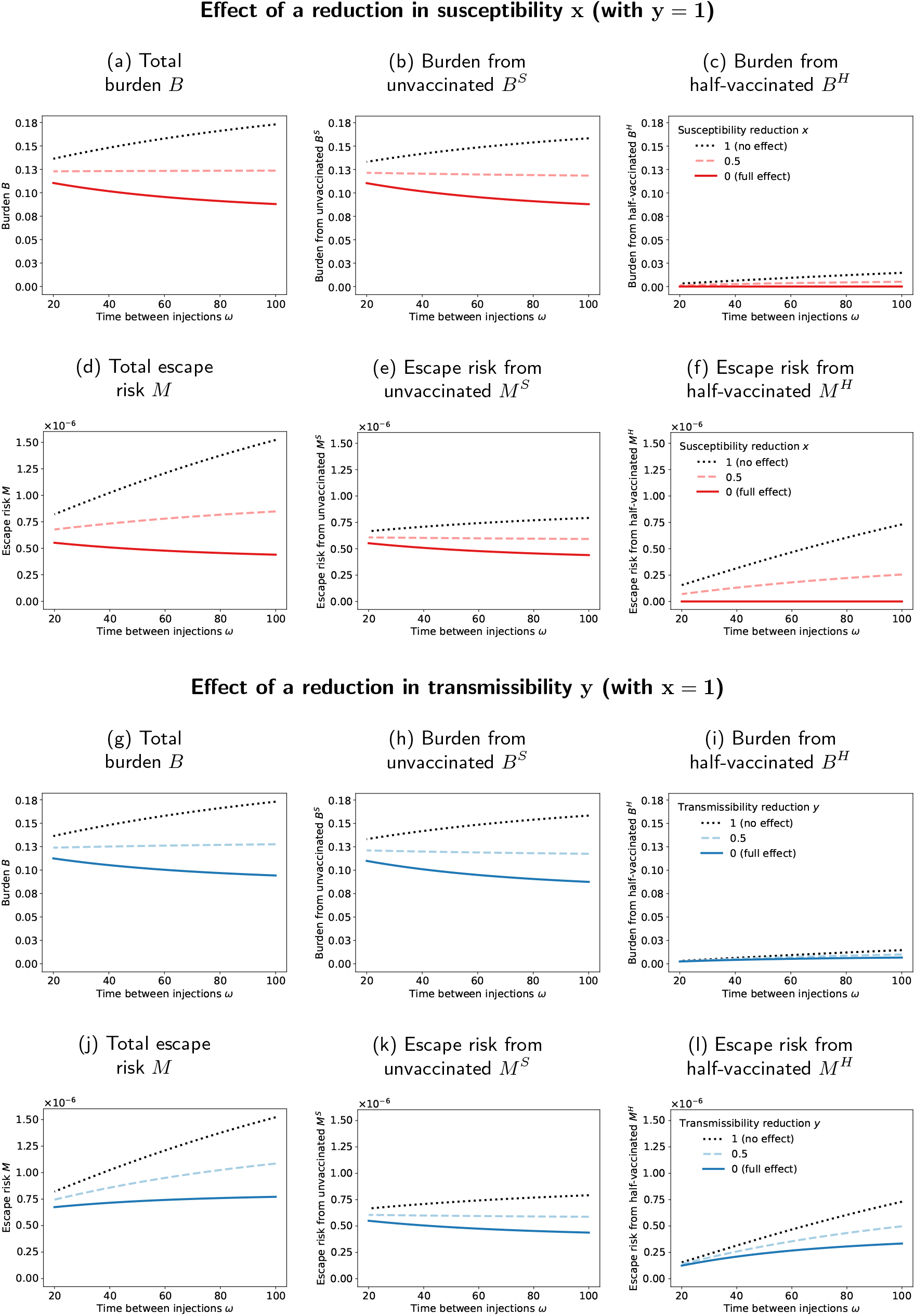
Burden and fraction of new mutant infections (escape risk) as a function of the prime-boost interval for the two limiting cases, in which the primer dose has an effect on susceptibility only (*y* = 1, Panels a-f) or on transmissibility only (*x* = 1, Panels g-l). The black dotted lines, where the primer dose has no effect at all (*x* = *y* = 1) are identical in the two parts of the figure. Reductions in susceptibility and transmissibility both reduce the burden and the risk of vaccine escape. The consequences of a delay of the booster shot depend on the effects of the primer dose. A delay may increase both the burden and the risk of vaccine escape (dotted black line in Panels a and d/Panels g and j), decrease both quantities (solid red line in Panels a and d), or decrease the burden at the cost of an inreased risk of vaccine escape (solid blue line in Panels g and j). The figure shows results for our default parameter set with *µ*_*V*_ */µ*_*I*_ = 10.

**Figure 3:**
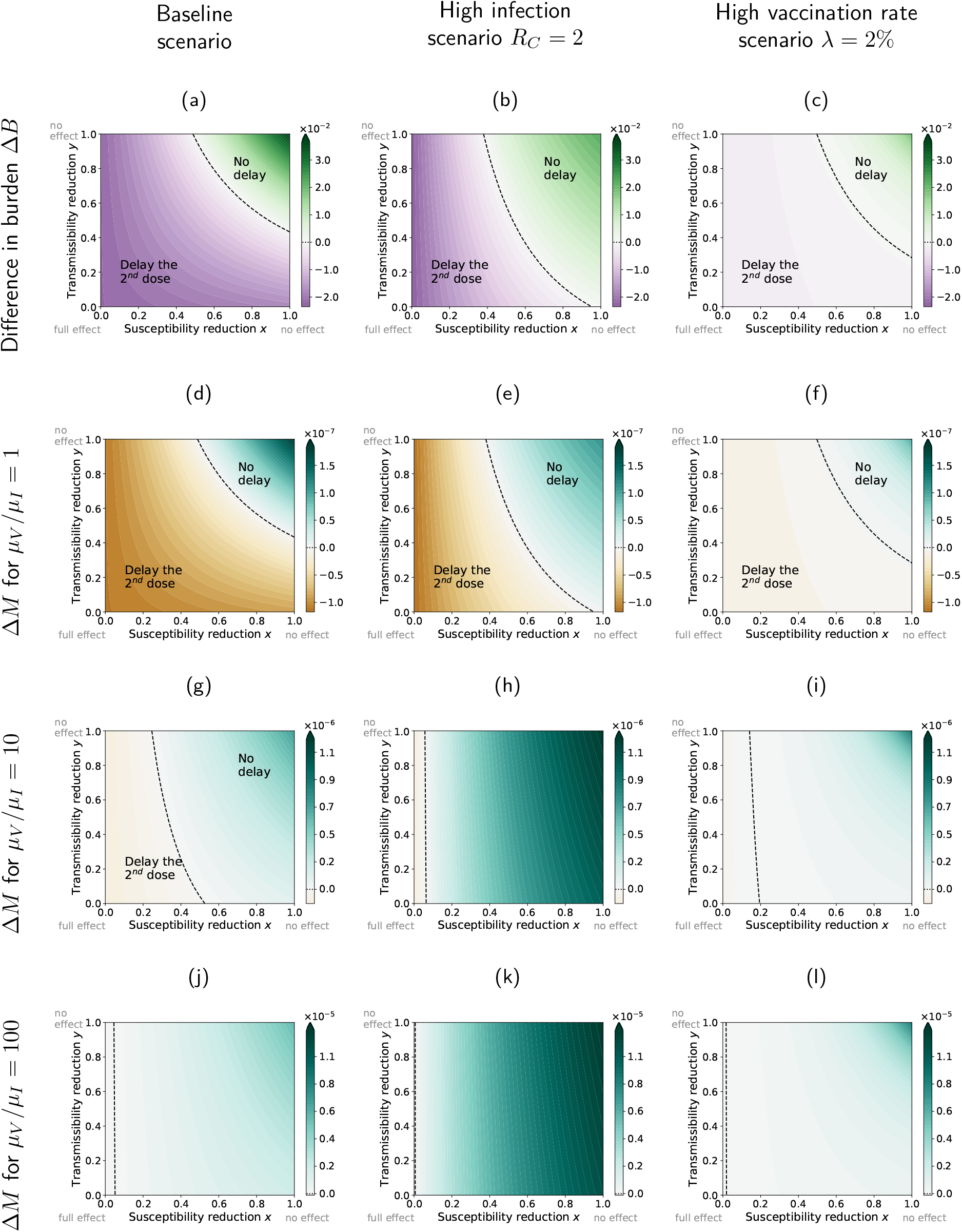
Differences in the burden and the fraction of new mutant infections between the strategies with a minimal and a maximal interval between the two vaccine doses, Δ*B* = *B*_*ω*=100_ *- B*_*ω*=20_ and Δ*M* = *M*_*ω*=100_ *- M*_*ω*=20_, depending on the reductions in susceptibility *x* and transmissibility *y* elicited by the primer dose. If the difference is positive, a delay increases the burden/escape risk (‘No delay’). If it is negative, a delay reduces the burden/escape risk (‘Delay the 2^nd^ dose’). The three columns correspond to our baseline scenario with the default parameter set (left column), a high infection scenario with a higher value of *R*_*C*_ (middle column), and a high vaccination rate scenario with a larger value of *λ* (right column). We consider three different ratios of the per-capita mutation rates in unvaccinated and half-vaccinated individuals. As expected, for *µ*_*V*_ */µ*_*I*_ = 1 (Panels d-f) the same strategy minimizes both the burden and the risk of vaccine escape, irrespective of *x* and *y*. For *µ*_*V*_ *> µ*_*I*_, there is a parameter range, in which a delay reduces the burden but increases the risk of vaccine escape.

In both cases, the burden mainly stems from unvaccinated individuals that become infected (compare *B*^*S*^ with *B*^*H*^) in Fig. 2. The prime-boost interval has not only a direct effect on this burden by affecting the fraction of unvaccinated individuals but also an indirect effect by changing the transmission dynamics (e.g. for *x* = 1, *B*^*S*^ increases with *ω*, although a longer delay reduces the fraction of unvaccinated susceptible individuals *S*). In a similar way, if the primer dose has no effect on transmissibility (*y* = 1), most mutants emerge from unvaccinated individuals. In contrast, if the primer dose has no effect on susceptibility (*x* = 1), evolution within half-vaccinated individuals substantially contributes to the emergence of vaccine escape even if the primer dose blocks transmission of the wild-type virus completely (*y* = 0, see Panel l).

Regarding the effect of a potential delay of the booster shot, we observe that the strategy *ω* that minimizes the burden is always either the no-delay strategy (*ω* = *ω*_min_) or a maximal delay (*ω* = *ω*_max_), but never an intermediate prime-boost interval (Panels a and g). The same holds true for the fraction of new mutant infections (Panels d and j). If the primer vaccination has neither an effect on susceptibility nor on transmissibility, both the burden and the escape risk increase with the time between the two injections. For strong effects in either susceptibility or transmissibility, an increase in the delay between injections reduces the burden (again, this is not true for higher *R*_*C*_, see below). For the escape risk, the picture is different. With a strong reduction in susceptibility, a delay of the booster shot reduces the escape risk. In that case, the effects of a delay on the burden and on the vaccine risk align. In contrast, if the vaccine has no effect on susceptibility (*x* = 1), delaying the second dose increases the risk of vaccine escape, irrespective of how well the primer dose blocks onward transmission of the wild-type virus (*y* = 0; solid curve in Panel j). The effects on the burden and on the risk of vaccine escape diverge in this case.

We therefore can conclude from these limiting cases that conflicts between reducing the burden and the risk of vaccine escape can exist (see *x* = 1, *y* = 0), but the reduction in cases can also outweigh the increased risk of within-host evolution of vaccine escape (see *y* = 1, *x* = 0).

### When do reductions in both transmissibility and susceptibility lead to trade-offs?

To investigate more closely under which circumstances the effect of the strategy on the burden and the fraction of escape mutants diverge, we proceed to explore the entire range of reductions in susceptibility and transmissibility and vary other parameters as well. As for the limiting cases, we found that the optimal strategy is either no delay or a maximal delay. We therefore focus on the burden and the escape risk with the recommended interval of *ω*_min_ = 20 days and a maximally stretched interval of *ω*_max_ = 100 days. To determine which strategy is optimal under the respective criterion, we consider the differences Δ*B* = *B*_*ω*=100_ *— B*_*ω*=20_ and Δ_*M*_ = *M*_*ω*=100_ *— M*_*ω*=20_ and ask when they are larger than zero (‘No delay’) or smaller than zero (‘Delay the 2^*nd*^ dose’), see Fig. 3. For reference, the absolute burden and fraction of new mutant infections with our default parameter set is given in Fig. 4 for both values of *ω*.

**Figure 4:**
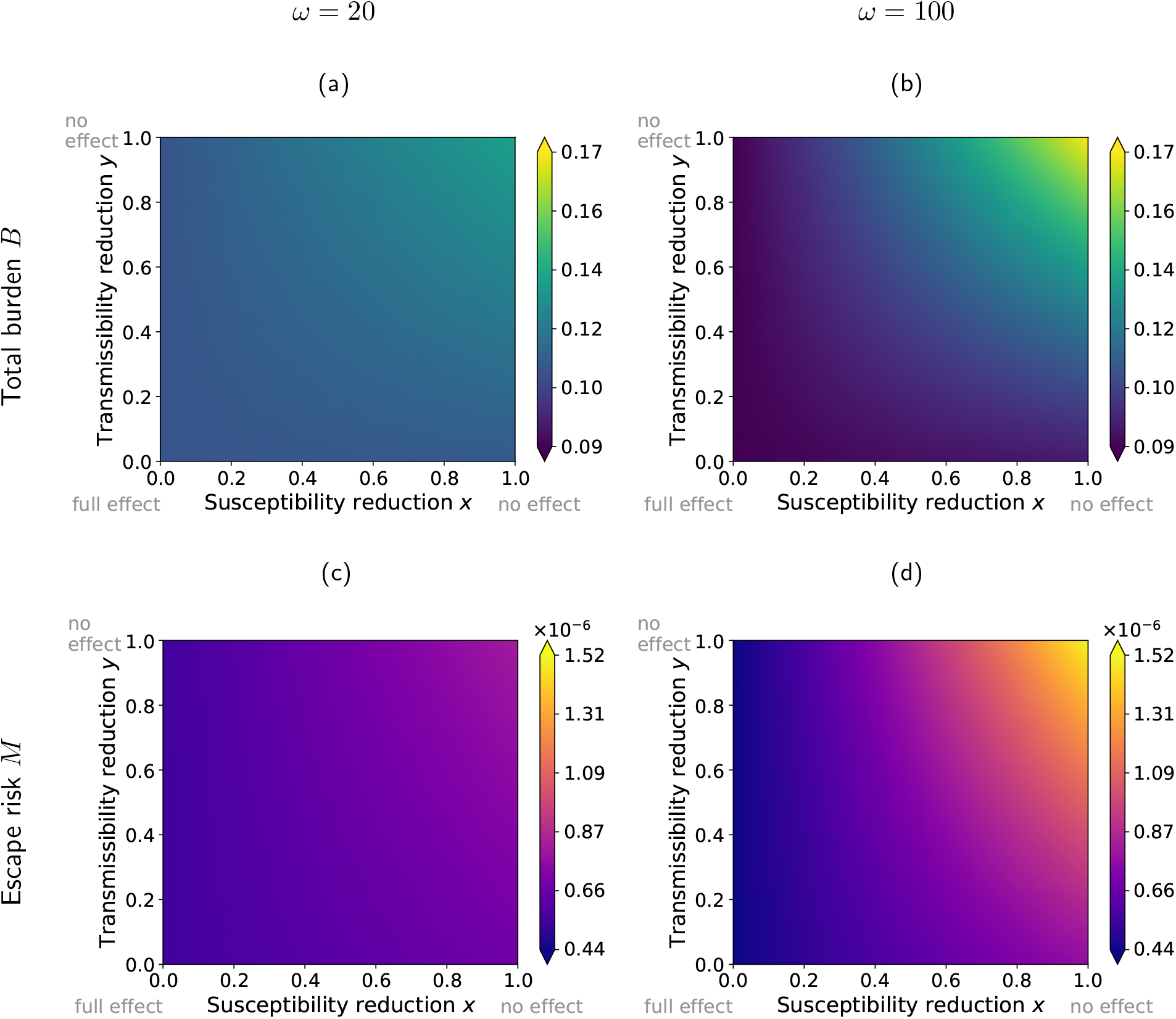
Total burden *B* and total number of new mutant infections *M* for a minimal prime-boost interval (Panels a and c) and for a maximal delay (Panels b and d), depending on the reductions in susceptibility *x* and transmissibility *y* elicited by the primer dose. Reducing the susceptibility or transmissibility reduces the total burden and the escape risk, but in quantitatively different ways such that a trade-off between them regarding the optimal *ω* can emerge. The figure shows results for our default parameter set with *µ*_*V*_ */µ*_*I*_ = 10.

We already know from the analysis of the limiting cases that neither strategy is optimal in minimizing the burden across the entire range of *x* and *y*. This can be further seen in Fig. 3a-c. If the effect of the primer dose is sufficiently strong, the delay strategy is favoured. If it is weak, the booster dose should not be delayed. The area in which delaying the booster dose is beneficial is reduced if *R*_*C*_ is higher, i.e. if the infection rate in the population is larger (cf. Panels a and b). In that case, a reduction of the transmissibility on its own without any effect of the primer dose on susceptibility is insufficient to justify a delay strategy. If more vaccine doses are available (higher vaccination rate *λ*, Panel c), the choice of the strategy becomes less important, with differences in outcomes being smaller.

Both strategies affect the burden and the fraction of escape mutants equivalently if *µ*_*V*_ = *µ*_*I*_ (compare the first two rows of the figure). For *µ*_*V*_ *> µ*_*I*_, as expected from the general considerations above, the range of *x* and *y* favoring a delay strategy is larger for the burden than for the number of mutants (Panels g-l). In that case, a parameter range opens up in which a delay strategy reduces the burden but increases the fraction of new mutant infections. Especially, if *µ*_*V*_ ≫ _*I*_, the risk of vaccine escape is increased by a delay strategy over nearly the entire range of *x* and *y* (Panels j-l).

The differences Δ*B* and Δ*M* between the two strategies can be substantial, such that they become relevant in choosing vaccination strategies. If vaccine doses are scarce, the strategy can change the infected fraction of the population by several percentage points of the population for some combinations of *x* and *y*. To put this into perspective, the cumulative fraction of Covid cases in Germany as of now (end of April) is around 7% (assuming as above that about 50% of all cases are detected (Backhaus et al.,2021)). Hence, the effect of the strategy is in some parameter regions of the same order of magnitude as the cumulative number of cases in Germany at the time this manuscript is written. Likewise, the strategy can substantially influence the total fraction of new mutant infections, doubling *M* in some cases and with it increasing the risk that vaccine escape mutants evolve.

### What changes if vaccines are available right from the start of the pandemic?

We finally compare the results to a scenario in which the vaccine is available right from the start of the pandemic, for which we choose *I*(0) = 10^*-*6^ and *R*(0) = 0 (Fig. 5). Such a scenario may be relevant in case escape mutations necessitate a novel vaccination campaign and that the vaccine is available before the new variant is present in all countries. In this case, the parameter range in which a delay strategy minimizes the risk of escape mutants is enlarged (compare Panels c and d). However, the benefit of either strategy in reducing the burden or the risk of vaccine escape is much smaller than for our primary scenario, in which a vaccine becomes available only during the pandemic (compare the two columns).

**Figure 5:**
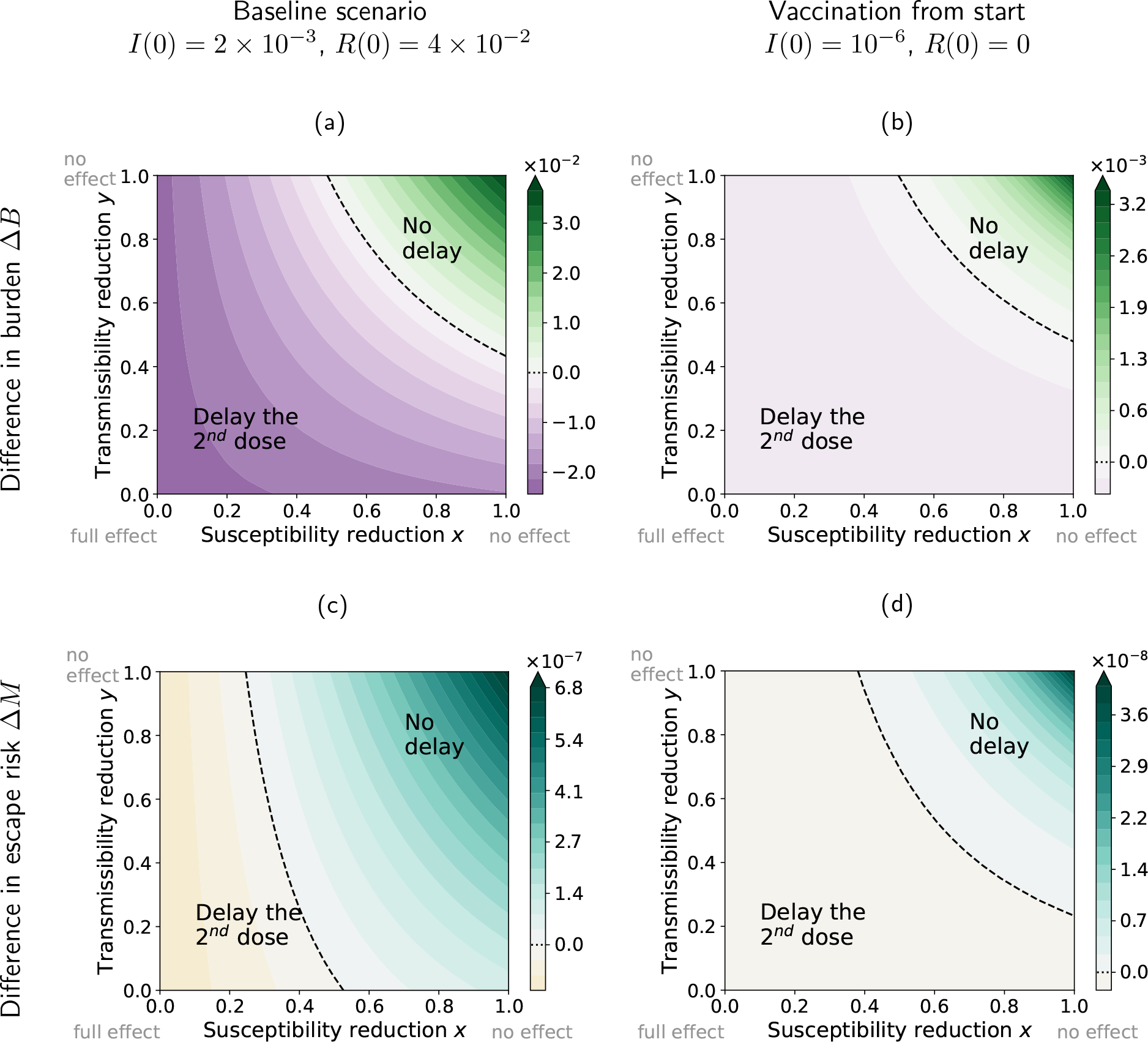
Comparison between a scenario, in which vaccines become available during a pandemic, (Panels a and c) and a scenario, where they are available right from the start (Panels b and d). The figure shows the differences in the burden and in the number of new mutants infections between the strategies with a minimal and a maximal interval between the two vaccine doses, Δ*B* = *B*_*ω*=100_*-B*_*ω*=20_ and Δ*M* = *M*_*ω*=100_ *- M*_*ω*=20_. When vaccines are available from the start, differences between strategies are much smaller. (Note the different scales of the color gradients.)

## Discussion

When vaccines are scarce, should we increase the timing between the primer dose and the booster dose? Using an SIR-model, extended by half-vaccinated (primer dose only) and fully vaccinated (primer+booster dose) individuals, we explored when the choice of the prime-boost interval – delay or no delay – leads to conflicts between reducing the burden and the risk of vaccine escape evolution. On short time scales, reducing the burden is the primary goal, but the risk of vaccine escape could pose a major societal problem on a longer time scale.

### What does the SIR model find?

If the primer dose has only a weak effect, a delay of the booster dose increases both the burden and the fraction of new mutant infections. If the primer dose has a sufficiently strong effect, a delay is beneficial in both respects. However, this latter area in the susceptibility-transmissibility reduction (*x*-*y*) plane is small and limited to an extremely high reduction in susceptibility if the within-host evolution of vaccine escape is much larger in half-vaccinated than in unvaccinated individuals. Between these two parameter ranges in which both criteria suggest the same prime-boost interval, there is a parameter region in which a delay reduces the burden – but at the cost of an increased risk of vaccine escape. This region is absent if vaccine escape evolution is equally likely within half-vaccinated and unvaccinated patients.

Our model suggests that the conditions under which a delay strategy is favorable for reducing the burden are more restrictive with a higher reproductive number *R*_*C*_. This is the opposite of what has been found by Matrajt et al.(2015), comparing a strict one-dose to a two-dose strategy – but not focussing on the scarcity of a vaccine. In addition, there are several differences between the models.

E.g. Matrajt et al.(2015) assume that all individuals are vaccinated at the same time but immunity takes time to build up following vaccination, while we assume that vaccine roll-out is a continuous process over time but once an individual has received a vaccine shot, the effect is immediate. Other differences include incomplete vs. complete vaccine coverage, presence and absence of asymptomatic infections, and incomplete vs. complete protection with two doses.

### What affects the fate of new escape variants?

We only consider the emergence of vaccine escape mutants but do not track their spread. As already mentioned in the model section, the fate of these mutants is subject to stochasticity as long as they are rare (Rella et al.,2021). Especially when the distribution of secondary cases is overdispersed – i.e. when a small number of patients infects many others, while the majority of patients infect only few or no other individuals –, it is likely that the mutation is lost again from the population (Lloyd-Smith et al., 2005). The heterogeneity in transmission has been estimated to be rather high for SARS-CoV-2 (Riou and Althaus,2020;Sun et al.,2021). Beyond these stochastic effects, there are many factors that affect the spread of escape variants by changing their reproductive number, and we only discuss a few examples here. Social distancing, contact tracing, and isolation of infecteds control not only the wild-type virus but also escape variants. For our model, we assumed *R*_*C*_ to be constant throughout the pandemic, but in reality the transmission coefficient changes over time due to control measures, changes in behaviour, and also up to some extent seasonality. This includes in particular the extent at which social distancing restrictions are lifted already during a vaccination campaign. This affects the establishment of escape variants (Rella et al.,2021), but also their propagation once frequent. A further factor that is crucial for the fate of escape mutants, short-term and long-term, is the degree up to which vaccination is still effective against them. Cobey et al.(2021) argue that vaccination will likely still grant some level of protection against escape variants. This means that they would not spread well in a population with high vaccination coverage. Yet, they may accumulate additional mutations over time that increase their degree of vaccine resistance. Apart from residual vaccine protection to escape mutants, if different vaccines using different antigenic targets or variants of the same target are employed, the spread of mutants across the population escaping from one vaccine is likely still hampered by the other vaccines (McLeod et al.,2021).

### Are effects on the burden and on vaccine escape equally predictable?

The difference that the choice of strategy makes can be substantial. Whether an extended interval between shots increases or decreases the burden, crucially depends on the reductions in susceptibility and transmissibility elicited by the primer dose. The number of studies estimating the effects of the primer (and booster) doses of the various Covid vaccines is currently rapidly growing. With sufficient information, we can probably be rather confident about the consequences of our choice with respect to the disease burden. Matters are much more complicated when it comes to assessing the risk of vaccine escape, which requires to make predictions about evolution in a highly complex and dynamic environment. In our simple model, the range of primer dose effects for which a reduction in burden comes at an increased risk of vaccine escape depends on the relative probabilities of within-host evolution of escape mutants in half-vaccinated and unvaccinated individuals. There is concern that vaccine escape mutants evolve more easily in partially immune individuals, but we do not know whether this is really the case and if so, how much more likely it is (Branswell,2021 Jan. 4;Cobey et al.,2021;Hanage and Russell,2021;Saad-Roy et al.,2021). We generally do not know how easily the virus can escape from immunity nor do we know up to which degree mutants will evade vaccine-induced immunity and potentially also naturally acquired immunity nor what their degree of cross resistance to other vaccines is going to be. And even if we knew all this, the appearance and establishment of vaccine escape mutants would still be a probabilistic event that may or may not happen. Moreover, escape mutants can also be imported from other regions (Gerrish et al.,2021). When sufficient information on the effectiveness of the primer dose and the duration of immunity is known, it is hence weighing an immediate assessable benefit against an unknown future risk with unknown consequences.

## Conclusions

Our model is not suitable to solve the dilemma nor to make any concrete recommendations. It only contains the most essential elements necessary to describe the epidemiological dynamics, which, of course, requires making many simplifying assumptions. The per-capita rates of escape evolution are model parameters, and the model explores the population-level consequences, given a certain ratio between the rates in half-vaccinated and unvaccinated patients. Such a fundamental model shows in a transparent manner how the verbal arguments that have been brought forward play out in a quantitative model. Maybe most importantly, the model allows to see that a delay strategy can increase or decrease the risk of vaccine escape and to identify when conflicts between reducing the burden and the risk of vaccine escape arise, where the effects of the primer dose in terms of reductions in susceptibility and transmissibility and the relative risks of vaccine escape in half-vaccinated and unvaccinated patients are key parameters. It could also provide a starting point for more detailed models that take further complications into account. We hope that our model helps to provide a more solid foundation for the discussion on vaccination strategies.

## Data Availability

The Jupyter notebook for running the model and reproducing the figures is available at: https://github.com/fgeoffroy/Vaccine-escape

## Acknowledgements

We thank Andrea Graham and Nikhil Sharma for helpful discussions. This project was made possible with financial support from the German Research Foundation (DFG) and was supported by the Research Training Group 2501 on Translational Evolutionary Research (RTG 2501 TransEvo).

## A Availability of booster doses

In this section, we show that there are at all times enough doses available to administer the second dose after the chosen time interval *ω*, as long as *S*(*t*) *>* 0, i.e. there never arises a situation in which the second dose needs to be further delayed due to vaccine shortage. For this, we need to show that 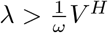.

We first consider a disease-free population 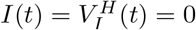. We denote the fraction of half-vaccinated individuals in this scenario by 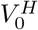. From Eq. (1), their dynamics is given by

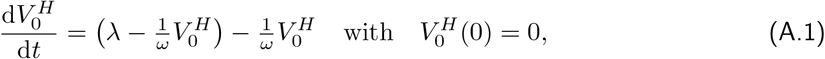

which solves to

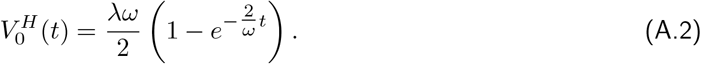

With this, we have:

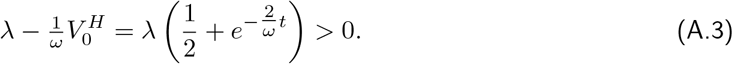

We now turn to a population, in which the virus is spreading. In this case, as given in Eq. (1), there is an infection in the differential equation describing the changes in the fraction of half-vaccinated individuals:

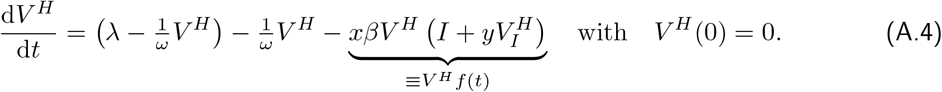

Comparing to Eq. (A.1), we see that

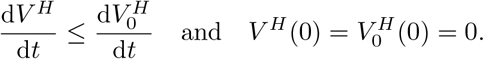

Therefore:

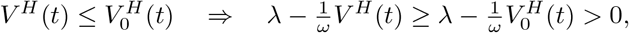

where the last step follows from Eq. (A.3). This implies 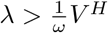.

